# Long-term impact of Training the Trainers program on Primary Eye Care networks in Malawi using the Arclight Project package

**DOI:** 10.64898/2026.03.31.26349901

**Authors:** Thomas Windle, Frank Maliko, Sarath Burgiss-Kasthala, Andrew Blaikie

## Abstract

**Background:** The World Health Organisation (WHO) advocates integrating primary eye care (PEC) within community health systems, supported by task-shifting and frugal technologies. While low-cost tools such as the Arclight device and Wilson anterior segment loupe have demonstrated training and diagnostic value, their long-term impact on community health worker (CHW) roles and professional networks remain poorly understood.

**Methods:** We conducted a qualitative follow-up study 3 years after implementation of an Arclight Project enabled cascade training of the trainers (ToT) PEC programme in central Malawi. Ophthalmic Clinical Officers (OCOs) trained using the Arclight training and diagnostic package subsequently cascaded PEC training to Health Surveillance Assistants (HSAs). Semi-structured interviews were undertaken 3 years later with OCOs and HSAs to explore device use, evolving professional roles, training diffusion, and communication patterns. Data were analysed thematically, informed by concepts from social network analysis to examine changes in advice-seeking, mentorship and peer collaboration.

**Results:** Frugal eye-care technologies functioned not only as diagnostic tools but as mechanisms of professional repositioning. HSAs equipped with low-cost diagnostic devices became recognised as community eye focal persons, receiving referrals from colleagues and community members. OCOs who delivered training emerged as central hubs for clinical advice and ongoing training, creating strong vertical networks between district and community levels. However, horizontal peer-to-peer networks among HSAs remained weak and largely informal. Communication relied heavily on ad-hoc phone calls and WhatsApp messaging, with limited structured communities of practice. Despite sustained use of devices and retention of key skills, network activity often declined over time without active reinforcement.

**Conclusions:** Frugal eye-care technologies act as social as well as clinical interventions, reshaping CHW networks and professional hierarchies. Designing PEC programmes with explicit attention to strengthening and sustaining professional networks, rather than focusing solely on skills transfer, may further enhance alignment with WHO Integrated People-Centred Eye Care and improve long-term programme sustainability and impact.

## Introduction

Access to quality eye care remains a major global public health challenge, particularly in low-and middle-income countries (LMICs), where over 90% of the world’s vision impairment occurs [1]. In response, the World Health Organisation (WHO) and the International Agency for the Prevention of Blindness (IAPB) have prioritised the integration of Primary Eye Care (PEC) into community health systems through the framework of Integrated People-Centred Eye Care (IPEC) [2,3]. Central to this approach is task sharing where eye care tasks are delivered by and shared between mid-level and community health workers to expand coverage improving equity of access strengthening health systems. However, despite strong policy commitment, implementing PEC at scale remains challenging constrained by workforce shortages, training limitations and lack of access to appropriate training and diagnostic technologies [4,5].

Frugal technologies aim to be low-cost and robust without compromising essential functionality. They are emerging as an important part of the solution to limited training and diagnostic capacity in low- and middle-income countries (LMICs) [6]. In eye and ear care an important example includes the Arclight Project package of training and diagnostic devices. The Arclight and Wilson devices are both anterior segment loupes and otoscopes with the Arclight also being a direct ophthalmoscope [7]. They are solar powered, highly portable, low-cost tools designed to enable identification of the most common anterior and posterior segment causes of eye disease found in LMICs [Fig 1] [7]. The package also includes near and distance visual acuity charts offering the opportunity for community and district level health care workers to deliver more comprehensive eye examinations as recommended by the WHO primary eye care manual and online training programmes [8–10]. These devices have been introduced into several LMICs where district-level clinicians were trained to subsequently train community-level health workers [11, 12]. While several studies have demonstrated short-term improvements in knowledge and examination skills following such interventions [11, 13, 14], far less is known about their long-term impact on health worker roles, relationships, and professional networks.

**Fig 1:**
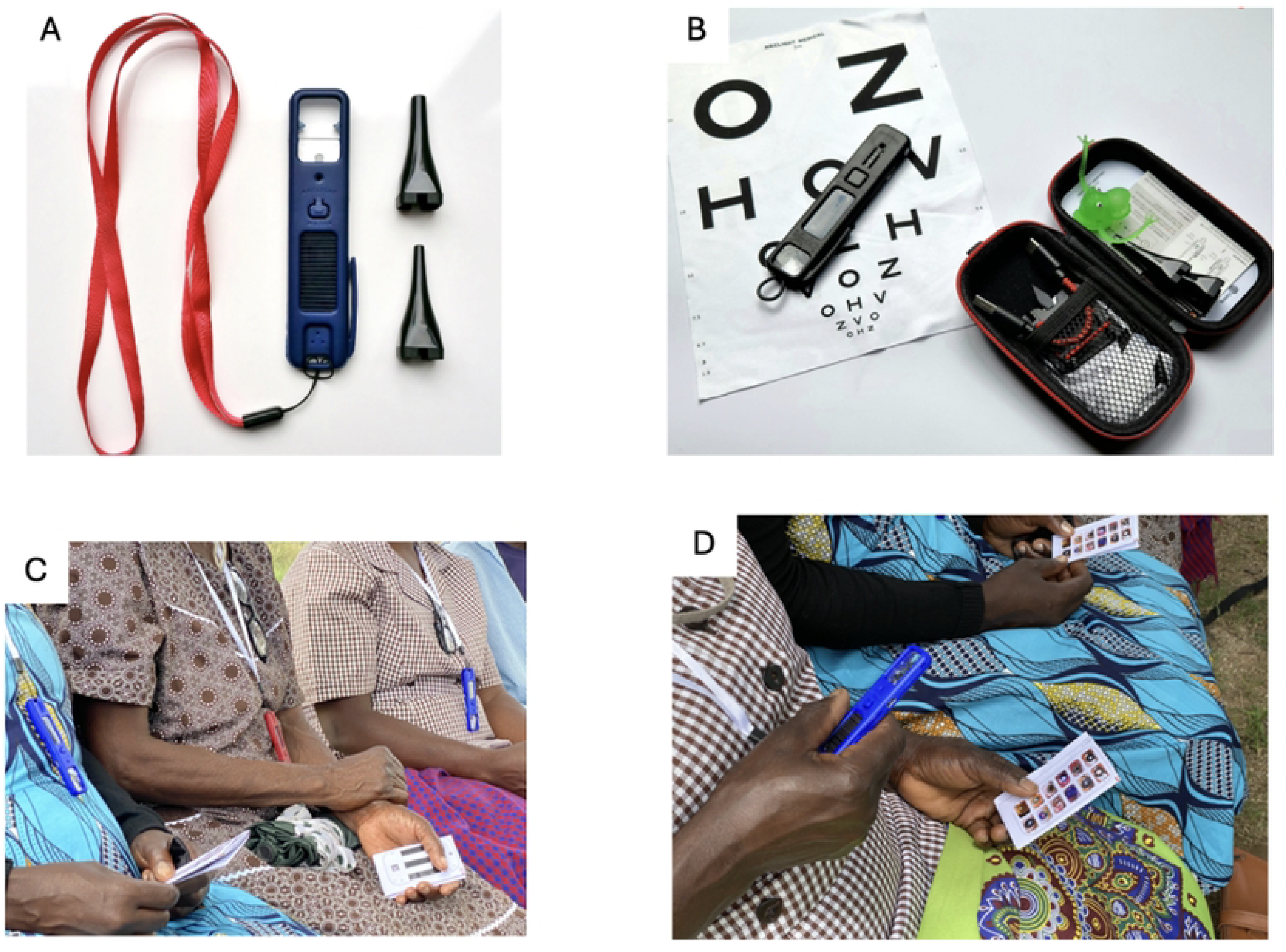
*(A)* The Wilson solar-powered anterior segment loupe *(B)* Contents of the Arclight device package, including the Arclight direct ophthalmoscope, loupe, otoscope, visual acuity chart and examination accessories. *(C–D)* Training of HSAs in anterior segment examination using Wilson loupe, illustrating hands-on skill acquisition for assessment of common causes of visual impairment in low-resource settings.

In Malawi, Health Surveillance Assistants (HSAs) form the foundation of community-level healthcare delivery, with responsibilities spanning maternal and child health, infectious disease control and health promotion [15]. Ophthalmic Clinical Officers (OCOs) provide district-level eye care services and supervision. In 2019–2020, a cascade PEC training programme was implemented in central Malawi, where OCOs were trained using the Arclight package of training and diagnostic tools and subsequently equipped and supported to train HSAs in core eye disease assessment and referral. Although initial internal programme evaluations suggested improvements in HSA knowledge and confidence, how these changes have been sustained over time and how they have influenced communication, supervision, and professional interaction between cadres, has not been explored.

Existing research on task-shifting in eye care has primarily focused on clinical competencies and service outputs, with limited attention to the social and relational dynamics that underpin sustainable health system change [16]. Health workers however do not operate in isolation. The ability to apply new skills depends heavily on access to advice, peer support, and supervision networks. Social network analysis offers a valuable means to examine how training interventions reshape patterns of communication and influence professional support within healthcare systems. Yet few studies in global eye health have applied this perspective, particularly using long-term qualitative follow-up [17].

This study seeks to address this gap by exploring how a frugal technology-enabled cascade PEC programme has influenced the roles and social networks of OCOs and HSAs in central Malawi, over three years after initial training. Through in-depth qualitative interviews analysed using social network methods, we examine the potential role of frugal technologies in professional identity, authority and inter-cadre relationships. By situating our findings within the WHO IPEC framework and current debates on task shifting and frugal innovation, we aim to contribute evidence on how primary eye care programmes can be designed for long-term sustainability, not only in skills retention but also in resilient professional networks [18–20].

## Methods

### Study design

This study used a qualitative design to explore the long-term social and professional impacts of a cascade primary eye care training programme in central Malawi. We conducted semi-structured in-depth interviews with Ophthalmic Clinical Officers (OCOs) and Health Surveillance Assistants (HSAs) in 2024 between September 16^th^ and October 11^th^. These health care workers had participated in a frugal technology enabled ToT cascade programme between 2019 and 2020. The study employed a thematic analysis approach, informed by concepts from social network analysis, to examine how professional roles, communication patterns and advice-seeking networks had evolved over time.

### Study setting

The study was conducted in selected districts of central Malawi where the primary eye care training programme had been implemented. Malawi’s health system is structured around a tiered primary healthcare model, with HSAs delivering community-level services and OCOs providing specialist eye care and supervision at district hospitals. The training programme equipped OCOs with frugal eye-care technologies, including the Arclight device and Wilson anterior segment loupe, and supported them to cascade training to HSAs within their catchment areas [Fig 2].

**Fig 2:**
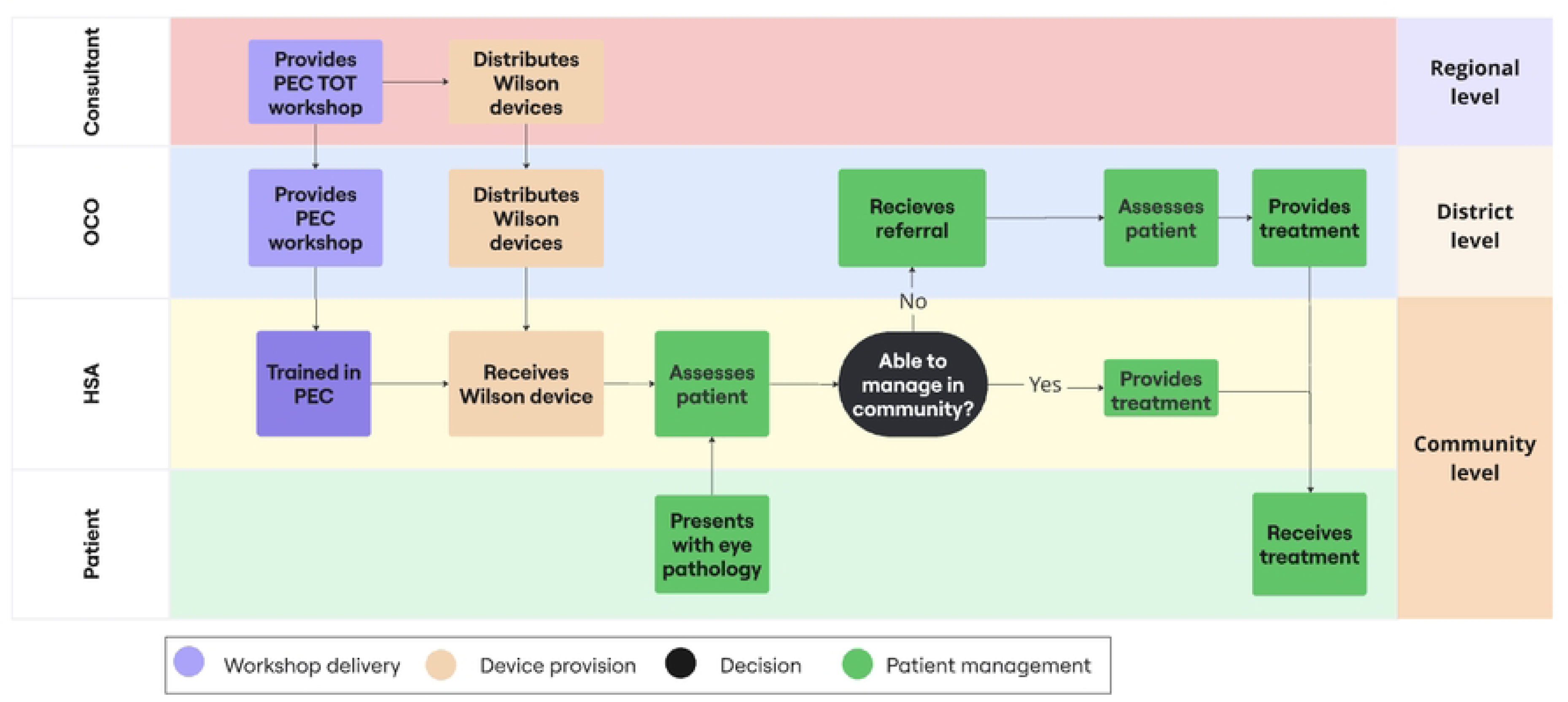
Swim-lane diagram depicting the cascade model for primary eye care (PEC) training in central Malawi. Ophthalmic Clinical Officers (OCOs) receive training to deliver district-level PEC workshops for Health Surveillance Assistants (HSAs) and to support the distribution of Wilson anterior segment loupes. Trained HSAs subsequently apply these skills in community settings to assess patients, manage uncomplicated eye conditions, and refer cases beyond their scope of practice to OCOs.

These districts were selected because they had completed at least three cycles of cascade training and had ongoing PEC activity at the time of data collection. The settings included a mix of rural and semi-urban communities, reflecting the diversity of service delivery environments in central Malawi.

### Participants and sampling

Two participant groups were included in the study:

1. Ophthalmic Clinical Officers (OCOs) who had completed the original ToT programme and subsequently trained HSAs.
2. Health Surveillance Assistants (HSAs) who had received PEC training from OCOs during the cascade phase.

Participants were purposively sampled to capture diversity in geography, years of post-training experience, and level of ongoing eye care activity. Selection criteria for OCOs included completion of ToT and active involvement in HSA training or supervision. HSAs were eligible if they had attended the PEC training and were still working in their communities at the time of study. A total of 9 OCOs and 8 HSAs were interviewed.

### Data collection

Data were collected between August 2025 to October 2025, approximately 3–4 years after the initial training intervention. Semi-structured interview guides were developed separately for OCOs and HSAs, covering topics such as:

- Use of frugal eye-care devices in daily practice
- Changes in professional roles and identity
- Experience of training others and receiving supervision
- Patterns of communication and advice-seeking
- Perceptions of support networks and collaboration

Interviews were conducted in English or Chichewa according to participant preference. All interviews were audio-recorded with participant consent and later transcribed verbatim.

Where interviews were conducted in Chichewa, transcripts were translated into English by bilingual members of the research team. Field notes were also taken during and after interviews to capture contextual observations and reflections.

### Data analysis

Data were analysed using a thematic analysis approach. Transcripts were read repeatedly to achieve familiarisation. An initial coding framework was developed both deductively (guided by the interview questions and study objectives) and inductively (allowing new themes to emerge from the data).

Coding was conducted manually and through qualitative data management software. Codes were grouped into broader categories and themes that reflected patterns in participants’ accounts. Analysis was further informed by concepts from social network analysis, including centrality, vertical and horizontal ties, and network density, to interpret how relationships and communication structures had evolved following the intervention.

### Ethical considerations

Ethical approval for this study was obtained from Nkhoma CCAP Hospital Research and Ethics Committee in Malawi and the University of Dundee Schools of Medicine and Life Sciences Research Ethics Committee.

All participants provided written informed consent prior to interview. They were informed of the purpose of the study, the voluntary nature of participation, and their right to withdraw at any point without consequence. All identifying information was removed from transcripts, and pseudonyms were used to ensure confidentiality. Audio files and transcripts were stored on password-protected devices accessible only to the research team.

## Results

This study draws on interviews with nine Ophthalmic Clinical Officers (OCOs) and eight Health Surveillance Assistants (HSAs). The findings are organised into five interlinked themes that describe how frugal devices, training and time reshaped professional roles and communication networks between cadres.

### 1. Frugal eye-care devices as tools and symbols of authority

Both OCOs and HSAs described how being given an Arclight device and/or Wilson anterior segment loupe improved positively their ability to assess patients and their perceived role within the health system.

> “I could identify a cataract easily compared to using naked eye. This meant I could diagnose more patients with cataracts.” (HSA, rural)

> “It is easier to decide what the problem is when on patient’s doorsteps rather than send them to the district hospital.” (HSA, rural).

HSAs frequently explained that possession of the device elevated their status within the health facility and community. Several participants noted that community members, colleagues and even other health workers began referring patients to them specifically for eye problems.

> “HSAs in the local area know that I have completed the PEC training, they will refer patients onto me as I am able to give them a better assessment.” (HSA, semi-rural)

Some HSAs described being referred to as “the eye doctor” within their communities despite not holding that formal role. The device acted not only as a clinical tool but as a visible symbol of specialised knowledge and authority.

> “Yes, it really helped us because people know this is a device to examine eyes and people now run to us to ask, “can you check my eyes”. We are known to be the doctors of the community on eye issues.” (HSA)

> “It has changed because now I feel like a doctor.” (HSA)

OCOs also described how the tools strengthened their ability to supervise and teach. They felt more confident demonstrating eye examinations and troubleshooting cases with HSAs during follow-up visits.

> “After attending the Train the Trainers workshop, I felt I had the knowledge and skills to train the HSAs. I was confident of doing the job of training them.” (OCO)

### 2. Emergence of vertical professional networks

Across interviews, strong vertical networks developed between OCOs and HSAs. HSAs commonly sought advice from OCOs regarding difficult cases, referral decisions and equipment use, particularly during the first years after training.

> “If I refer a patient, I will always call an OCO first to learn more about the condition I am referring. I also received an award at the district hospital for being a HSA that referred the most patients with eye problems which was nice.”

Most HSAs described OCOs as their primary point of contact for clinical reassurance.

> “I always go to the OCOs at the district hospital. When I go to a district hospital, the OCO will explain that a condition is called this and the treatment is this. Point of referral is at district hospital level so this is where advice comes from.” (HSA)

> “The HSAs call us for advice, and we act like a consultant for them.” (OCO)

OCOs confirmed that they regularly received calls and messages from multiple HSAs, often acting as intermediaries between community-level health workers and ophthalmologists or district hospital services.

> “I have HSAs calling me every day to ask me about things they are not sure about. For example, we had an issue with pink eye, they were asking me about this and how to manage it.” (OCO)

This created a hierarchical communication pattern, where most advice flowed vertically upwards rather than laterally across the HSA peer group.

### 3. Weak horizontal peer networks among HSAs

While vertical links were strong, lateral peer-to-peer networks among HSAs were much less developed. Most HSAs reported limited regular interaction with fellow trained HSAs regarding eye care, beyond occasional informal discussions or during workshops.

Several participants described only interacting with other trained HSAs during initial training or rare refresher sessions.

> “No, there hasn’t been much interaction as we only met once during the workshop”. (HSA)

Even when multiple trained HSAs worked within the same district, there were few structured platforms for case discussion or shared learning. As a result, collaboration tended to depend on individual motivation rather than organised systems.

OCOs showed more promising peer-to-peer networks with many stating that they will call OCO colleagues for advice before escalating to specialist care services.

> “If the issue can be handled by a fellow OCO, I would first ask them.” (OCO).

> “We met different OCOs from different areas which was helpful. After the training we were able to follow up with each other.” (OCO)

> “I feel that before the TOT workshop, I wouldn’t really sit down with colleagues to discuss things. But now we are all doing better at PEC as we discuss things more often.” (OCO).

### 4. Communication patterns and digital connectivity

Most participants described using mobile phones for communication about eye care. Phone calls and WhatsApp messaging were commonly used between HSAs and OCOs, particularly for urgent advice or referral decisions.

However, communication was largely ad hoc and unstructured. There were no formalised case discussion groups or ongoing communities of practice sustained by the programme.

“We have each other’s numbers, but there is no official group for eye cases. It is just when you need help.” (OCO)

Although digital tools were widely present, participants reported that workload, airtime costs and lack of structured facilitation limited their consistent use for professional learning.

“It would be useful to have an application we can have on our phones with photos of the fundus and back of the eye to show examples to HSAs of conditions.” (OCO).

### 5. Long-term retention of skills with gradual network decay

Most participants reported they still used the Arclight device and Wilson loupe several years after training, particularly for screening patients with red eye, blurry vision and suspected cataract. However, patterns of consultation and collaboration had reduced over time.

HSAs described their confidence remaining higher than before training, but some reported less frequent engagement with trainers as time passed.

OCOs also reported that communication intensity had declined, often due to increasing workload, staff transfers/deaths and lack of refresher activities from the programme.

> “I think the content overall was ok. The limiting factor for me was just having one session. Further training sessions would be very welcome. (OCO).”

Despite this decline, participants felt the programme had a lasting impact on their practice and professional identity. However, many emphasised that without ongoing support and structured interaction, some of the earlier network momentum was difficult to sustain.

> “We want to have refreshers. Regular refresher sessions would be good.”

## Discussion

This study explored the long-term social and professional effects of a frugal technology enabled cascade primary eye care (PEC) training programme in central Malawi. We focused on how training and equipping with the Arclight and Wilson low-cost solar powered diagnostic tools can influence community health worker roles and networks over a period of three to four years. Our findings demonstrate that the devices and associated training acted not only as diagnostic tools but also as social catalysts, influencing professional identity, hierarchies, and patterns of communication between cadres. These effects are highly relevant to the current WHO and IAPB agenda on Integrated People-Centred Eye Care (IPEC), where sustainable scale-up depends not only on skills transfer but on strengthening health-worker systems and relationships [2].

### Frugal technology as a social intervention

Much of the literature on frugal health innovation has focused on affordability, portability and diagnostic accuracy [6]. Our study adds to this by demonstrating that devices such as the Arclight device and Wilson anterior segment loupe also function as symbolic and relational tools. For HSAs, possession of the device conferred professional legitimacy, authority and recognition within communities and health facilities. Participants described becoming known as the “eye doctor”, with increased referrals from colleagues and community members.

This suggests that diagnostic tools should be understood to not only be that but also as interventions that reshape professional identities and social positioning. In settings where community trust and legitimacy are central to service utilisation, this symbolic dimension may be as important as technical performance.

### Task shifting, networks and IPEC

WHO’s IPEC strategy emphasises integration, teamwork and people-centred care across all levels of the health system [20]. Task shifting from specialist eye care staff to mid-level and community-based workers is essential for achieving universal eye health coverage in low-resource settings. However, most task-shifting programmes focus on individual competencies and service outputs, rather than the social systems that enable cadres to function effectively [20, 21].

Our findings highlight that task shifting in PEC does not just give the individual new skills and responsibilities in isolation but also impacts positively on strengthening professional networks. Vertical ties between OCOs and HSAs became stronger with OCOs acting as central hubs for supervision and advice facilitating clinical support and referral flow.

From an IPEC perspective, this hub and spoke model presents both an opportunity and a vulnerability. Strong vertical ties are beneficial for supervision, quality control and escalation of complex cases. However, over-reliance on a few OCOs risks collapse of networks if they are transferred, overburdened or leave the system. Designing task-shifting interventions that deliberately foster both vertical and horizontal ties may enhance resilience and sustainability [21].

### Weak horizontal networks and implications for sustainability

Despite strong vertical relationships, horizontal peer networks among HSAs remained weak and informal. There was little evidence of sustained peer-to-peer learning or structured communities of practice, even several years post-training. This pattern is consistent with broader literature on health worker training in LMICs, where initial enthusiasm often declines without deliberate mechanisms for ongoing engagement [21,22,23]. A systematic scoping review of ongoing training for CHWs in LMICs found significant variability in design and delivery of programmes, with many focusing on initial or refresher skills rather than structured, sustained mechanisms to support ongoing learning, supervision or peer networks. This highlights a gap in understanding how best to maintain competencies long term [24].

The limited lateral networking observed suggests a missed opportunity. Communities of practice which can be informal or formal groups where practitioners share experiences, reflect on challenges and support one another are increasingly recognised as important for sustaining behaviour change and professional development. But systematic reviews have shown that these communities vary considerably in the way they are structured and operate [25]. Incorporating structured peer learning platforms, such as moderated WhatsApp groups or periodic online learning sessions, could strengthen these lateral ties and reduce reliance on hierarchical support alone.

### Long-term follow-up

A strength of this study is the long-term qualitative follow-up, conducted three to four years after the initial training. Many task-shifting studies assess outcomes immediately or within months after intervention with few examining how relationships, identities and practices evolve over several years. Our findings suggest that while technical skills and device use were largely retained and new networks were established these declined over time without deliberate reinforcement.

This temporal perspective is critical for global eye health policy and programming. It indicates that sustainability depends not only on the initial training model with distribution of diagnostic tools but on also putting into place ongoing mechanisms such as refresher training, mentorship and where possible digital communication platforms.

### Implications for programme design and policy

These findings highlight two key implications for the design of future primary eye care programmes. First, training initiatives should be designed to strengthen professional networks, not just individual skills, by actively incorporating mechanisms for mentorship and peer-to-peer learning that help sustain practice over time. Second, ownership of frugal technologies such as the Arclight plays an important role in positively reshaping how community health workers perceive themselves and how they are perceived by others, supporting increased eye-care activity and strengthening patient trust.

### Strengths and limitations

The main strength of this study lies in its long-term follow-up and application of a social network perspective to analyse PEC training. This is an approach that is rarely used in global eye health research. By capturing both HSA and OCO perspectives, we were able to analyse relationships across health system levels.

However, the study has several limitations. The sample size was small and geographically limited to central Malawi, which may affect transferability to other settings. As with all qualitative research, findings are based on self-reported experiences and may be influenced by recall or social desirability bias. There was risk of observer bias as the individuals supporting interview translation were also involved in the training of OCOs and HSAs.

Nevertheless, the depth of interviews allowed for rich insights into processes often missed by quantitative evaluations.

## Conclusion

This study demonstrates that frugal eye-care technologies operate not only as clinical tools but as catalysts for professional network development and identity transformation among community health workers. For WHO and IAPB’s vision of integrated, people-centred eye care to be realised, future programmes must move beyond skills acquisition alone and but also actively cultivate networks, relationships and communities of practice that sustain task-shifted eye care in the long term.

## Data Availability

Due to the qualitative nature of this study and the risk of participant identification within a small professional network, full interview transcripts cannot be made publicly available. De-identified excerpts supporting the findings are included within the manuscript. Additional de-identified data may be made available upon reasonable request to the corresponding author, subject to approval by the relevant institutional ethics committees.

## Author contributions

^1^Conceptualization, methodology, data curation, formal analysis, project administration, visualisation, writing – original draft preparation, writing – review & editing.

^2^Project administration, investigation.

^3^Formal analysis.

^4^Conceptualization, methodology, formal analysis, supervision, visualization, writing – review & editing.

## References

1. World Health Organization. Eye care in health systems: guide for action [Internet]. Geneva: World Health Organization; 2022 [cited 2025 Dec 7]. Available from: https://www.who.int/publications/i/item/9789240050068

2. International Agency for the Prevention of Blindness. Integrated people-centred eye care: advocacy to action toolkit [Internet]. London: IAPB; 2022 [cited 2025 Dec 7]. Available from: https://www.iapb.org/learn/resources/integrated-people-centred-eye-care-advocacy-to-action-toolkit/

3. Gupta N, Manna S, Senjam SS, Gupta V, Vashist P. Integrating primary eye care with primary health care: tracing the journey. Community Eye Health J. 2021;34(113):S5–S6. Available from: https://cehjournal.org/articles/202

4. Kalua, K., Gichangi, M., Barassa, E., Eliah, E., Lewallen, S., & Courtright, P. (2014). A randomised controlled trial to investigate effects of enhanced supervision on primary eye care services at health centres in Kenya, Malawi and Tanzania. BMC Health Services Research, 14(S1), S6. 10.1186/1472-6963-14-S1-S6

5. Somerville, J. G., Strang, N. C., & Jonuscheit, S. (2024). Task-shifting and the recruitment and retention of eye care workers in under-served areas: a qualitative study of optometrists’ motivation in Ghana and Scotland. Primary Health Care Research & Development, 25, e30. 10.1017/S1463423624000185

6. Weyrauch T, Herstatt C. What is frugal innovation? Three defining criteria. J Frugal Innov. 2017;2:1. doi:10.1186/s40669-016-0005-y.

7. Blaikie, A., Sandford-Smith, J., Tuteja, S. Y., Williams, C. D., & O’Callaghan, C. (2016). Arclight: a pocket ophthalmoscope for the 21st century. BMJ, i6637. 10.1136/bmj.i6637

8. 8- Malik, A. N. J., Mnedeme, G. F., Iriya, N., Bahati, P., Marealle, H., Blaikie, A., & Mafwiri, M. (2025). Evaluation of primary healthcare worker training to screen children under 5 years of age with a low-cost alternative to the direct ophthalmoscope, the ‘Arclight’, as part of the Integrated Management of Newborn and Childhood Illness (IMNCI) programme in Tanzania. BMJ Paediatrics Open, 9(1), e003520. 10.1136/bmjpo-2025-003520

9. World Health Organization Regional Office for Africa. Primary eye care training manual: a course to strengthen the capacity of health personnel to manage eye patients at primary-level health facilities in the African Region. Brazzaville: WHO Regional Office for Africa; 2018. Available from: https://iris.who.int/handle/10665/272970

10. Learning on TAP. Eye and vision care – Learning on TAP [Internet]. GATE-TAP; 2025 [cited 2026 Feb 2]. Available from: https://www.gate-tap.org/course-list/eye-and-vision-care/

11. Blundell, R., Roberts, D., Fioratou, E., Abraham, C., Msosa, J., Chirambo, T., & Blaikie, A. (2018). Comparative evaluation of a novel solar powered low-cost ophthalmoscope (Arclight) by eye healthcare workers in Malawi. BMJ Innovations, 4(2), 98–102. 10.1136/bmjinnov-2017-000225

12. Mndeme FG, Mmbaga BT, Kim MJ, Sinke L, Allen L, Mgaya E, et al. Red reflex examination in reproductive and child health clinics for early detection of paediatric cataract and ocular media disorders: cross-sectional diagnostic accuracy and feasibility studies from Kilimanjaro, Tanzania. Eye (Lond). 2021 May;35(5):1347–1353. doi:10.1038/s41433-020-1019-5.

13. Gore, R., Wang, J. N., Yang, C. D., An, M., Hunter, S. C., Shahraki, K., Blaikie, A., & Suh, D. W. (2025). Effective low-cost pediatric vision screening by naive nonophthalmic examiners using the ‘Arclight’ device. Indian Journal of Ophthalmology, 73(1), 41–44. 10.4103/IJO.IJO_3027_23

14. Tuteja, S. Y., Blaikie, A., & Kekunnaya, R. (2021). Identification of amblyogenic risk factors with the Brückner reflex test using the low-cost Arclight direct ophthalmoscope. Eye, 35(11), 3007–3011. 10.1038/s41433-020-01341-9

15. Kok, M. C., Namakhoma, I., Nyirenda, L., Chikaphupha, K., Broerse, J. E. W., Dieleman, M., Taegtmeyer, M., & Theobald, S. (2016). Health surveillance assistants as intermediates between the community and health sector in Malawi: exploring how relationships influence performance. BMC Health Services Research, 16(1), 164. 10.1186/s12913-016-1402-x

16. Okwen, M., Lewallen, S., & Courtright, P. (2014). Primary eye care skills scores for health workers in routine and enhanced supervision settings. Public Health, 128(1), 96–100. 10.1016/j.puhe.2013.10.007

17. Sabot, K., Wickremasinghe, D., Blanchet, K., Avan, B., & Schellenberg, J. (2017). Use of social network analysis methods to study professional advice and performance among healthcare providers: a systematic review. Systematic Reviews, 6(1), 208. 10.1186/s13643-017-0597-1

18. Lee, L., Moo, E., Angelopoulos, T., Dodson, S., & Yashadhana, A. (2023). Integrating eye care in low-income and middle-income settings: a scoping review. BMJ Open, 13(5), e068348. 10.1136/bmjopen-2022-068348

19. Yasmin, S., & Schmidt, E. (2022). Primary eye care: opportunities for health system strengthening and improved access to services. International Health, 14(Supplement_1), i37–i40. 10.1093/inthealth/ihab062

20. World Health Organization. World Report on Vision. Geneva: WHO; 2019.

21. Schneider H, Lehmann U. From community health workers to community health systems: time to widen the horizon? Health Syst Reform. 2016;2(2):112–118.

22. Kok MC, Dieleman M, Taegtmeyer M, Broerse JEW, Kane SS, Ormel H, et al. Which intervention design factors influence performance of community health workers in low- and middle-income countries? A systematic review. Health Policy Plan. 2015;30(9):1207–1227.doi:10.1093/heapol/czu126.

23. Hill Z, Dumbaugh M, Benton L, Källander K, Strachan D, ten Asbroek AHA, et al. Supervising community health workers in low-income countries—a review of impact and implementation issues. Glob Health Action. 2014;7:24085.doi:10.3402/gha.v7.24085.

24. O’Donovan J, O’Donovan C, Kuhn I, Sachs SE, Winters N. Ongoing training of community health workers in low-income and middle-income countries: a systematic scoping review of the literature. BMJ Open. 2018 Apr 28;8(4):e021467. doi: 10.1136/bmjopen-2017-021467.

25. Ranmuthugala G, Plumb JJ, Cunningham FC, Georgiou A, Westbrook JI, Braithwaite J. How and why are communities of practice established in the healthcare sector? A systematic review of the literature. BMC Health Serv Res. 2011 Oct 14;11:273. doi: 10.1186/1472-6963-11-273.

